# T2DFitTailor: A tool for type 2 diabetes patients to tailor exercise plan

**DOI:** 10.1101/2024.03.04.24303717

**Authors:** Cheng Liu, Xinyu Hou, Bingxiang Xu, Tiemin Liu, Ru Wang

## Abstract

Type 2 Diabetes (T2D) poses a formidable global health challenge, with its escalating prevalence contributing significantly to increased morbidity, mortality, and healthcare costs. This chronic condition, characterized by insulin resistance and hyperglycemia, leads to a plethora of complications, intensifying its societal and economic impact. Exercise, as a fundamental element of T2D management, is universally recommended for its ability to improve glycemic control and mitigate cardiovascular risks. However, individual responses to exercise vary considerably among T2D patients, leading to inconsistencies in the effectiveness of generic exercise plans. This heterogeneity in response to different exercise interventions highlights the need for more personalized approaches, tailoring exercise plans to individual patient profiles to optimize therapeutic outcomes. Our study addresses this critical need through the development of the T2DFitTailor package, a novel R-based tool that customizes exercise recommendations for T2D patients. Utilizing a comprehensive analysis of individual parameters, T2DFitTailor accurately predicts the impact of specific exercise plans on the reduction of glycated hemoglobin (HbA1c) levels. This personalized approach to exercise prescription in T2D management is a significant step forward in optimizing treatment strategies. It allows for a more targeted, effective intervention, thereby improving patient outcomes and quality of life. T2DFitTailor, accessible at ‘https://cran.r-project.org/package=T2DFitTailor’, represents a notable contribution to T2D care, reflecting the ongoing shift towards more data-driven and personalized healthcare strategies. This tool provides a practical solution to meet the varied requirements of T2D patients, ensuring that exercise prescriptions are not only based on scientific evidence but also customized to suit individual health profiles and specific needs.

## Introduction

Type 2 Diabetes (T2D) poses a significant global health dilemma. Global Burden of Diseases, Injuries, and Risk Factors Study (GBD) 2021 Diabetes Collaborators anticipate that by the year 2050, the global prevalence of diabetes will affect over 1.31billion people, with over 90% diagnosed with T2D,^(1)^ underscoring the urgency for effective management strategies. Characterized by insulin resistance and chronic hyperglycemia, ^(2)^ T2D contributes to a substantial increase in morbidity and mortality, inflicting considerable societal and economic burdens. The complexity of its management is further amplified by the disease’s multifactorial nature, necessitating a multifaceted approach to treatment.^(3),(4),(5)^

Among the key strategies for T2D management, exercise constitutes a cornerstone intervention, endorsed universally for its beneficial effects on glucose regulation and cardiovascular health.^(6),(7)^ However, a growing body of evidence reveals considerable variability in individual responses to specific forms of exercise (reference),^(8),(9),(10)^ suggesting a one-size-fits-all approach may not be optimal. This heterogeneity in glycemic response underscores the necessity for personalized exercise prescriptions that tailor intervention strategies to individual profile, a need scarcely addressed by existing tools and methodologies.

To address this pivotal need, we developed the T2DFitTailor package, a novel resource designed to tailor personalized exercise plans for patients with T2D, based on a comprehensive analysis of individual patient parameters. This tool not only facilitates the customization of exercise interventions but also predicts subsequent reductions in HbA1c levels to enhance glycemic control, thereby representing a significant advancement in the personalized management of T2D. The T2DFitTailor package is readily accessible at “https://cran.r-project.org/package=T2DFitTailor”, marking a pivotal step towards optimizing diabetes care through tailored exercise planning.

## Methods

### Data Resources

Our research utilized an extensive data collection process, focusing on the effects of exercise on Type 2 Diabetes (T2D). The search for relevant data was conducted through the PubMed database, employing a comprehensive set of keywords to ensure the inclusion of pertinent studies. The keywords used for this search were (“Type 2 Diabetes” OR “Diabetes Mellitus Type 2” OR “T2DM”) AND (“Exercise” OR “Physical Activity” OR “Aerobic Training” OR “Resistance Training” OR “Exercise Therapy” OR “Physical Fitness” OR “Physical Exercise”). The raw data are available at supplementary materials.^(11),(12),(13)^

### Definition of Responder and Non-Responder

In our analysis, individuals were categorized based on their HbA1c response to the exercise interventions. A sample was defined as a responder if there was a positive reduction in HbA1c levels following the exercise intervention. Conversely, a sample was categorized as a non-responder if no change or an increase in HbA1c levels was observed post-intervention.^(10)^

### LASSO Regression Analysis

Least Absolute Shrinkage and Selection Operator (LASSO) regression, a variation of linear regression, employs shrinkage to promote model simplicity and sparsity by driving some regression coefficients towards zero. This approach effectively eliminates variables with null coefficients, simplifying the model selection process and mitigating overfitting. In our research, LASSO regression was instrumental in isolating pivotal features relevant to HbA1c fluctuation post-exercise intervention. We implemented LASSO regression in Python using the “sklearn.linear_model” module, with the function of “Lasso”.

### Leave-One-Out (LOO)

Leave-One-Out (LOO) cross-validation, a technique where each observation in the dataset is sequentially used as the validation data while the rest serve as the training set, offers a comprehensive assessment of a regression model’s predictive accuracy, particularly beneficial in contexts of limited data size. In our study on Type 2 Diabetes (T2D), LOO was crucial for validating the precision of our regression model in predicting HbA1c reduction following exercise interventions. This method enabled a close examination of how well the predicted HbA1c reductions mirrored actual outcomes, ensuring robustness and mitigating overfitting risks. Our implementation of LOO, aligned with the project’s aim to measure the effectiveness of personalized exercise plans in managing T2D, employed the function of “LeaveOneOut” in python’s “sklearn.model_selection” module.

### K-Nearest Neighbors (KNN) Algorithm

The k-Nearest Neighbors (kNN) algorithm, a fundamental machine learning method for both classification and regression tasks, operates on the principle of utilizing the nearest data points in the feature space to predict the outcome for a new sample. In our research focused on Type 2 Diabetes (T2D), we employed the kNN algorithm to predict the HbA1c reduction in patients following various exercise interventions, identifying them as either responders or non-responders based on their individual profile variation. To fine-tune our model, we determined the optimal ‘k’ value through Leave-One-Out (LOO) cross-validation on the training set, ensuring a balance between bias and variance that is critical for the predictive accuracy of the model. Our analysis found that different exercise plan required distinct ‘k’ values for optimal results, with Taiji and Cycling performing best at k=2, Qigong, Stretching, and Walking at k=5, and Rugby at k=4. This LOO process not only facilitated the identification of the best ‘k’ values but also verified the model’s effectiveness on the training set. We further validated our model against a preserved test set based on the whole training set with above k values. The finalized kNN models on T2DFitTailor were built on the combination of training and test set with optimal k values. The implementation of the kNN regression was achieved utilizing the “sklearn.neighbors module” module with the function of “KNeighborsRegressor”.

## RESULTS

### Tailored exercise plan from T2DFitTailor

In this section, we elucidate the application of the T2DFitTailor package, which is designed to develop personalized exercise plans for individuals with Type 2 Diabetes (T2D) and forecast the HbA1c outcomes of these plans. The process requires inputting seven essential parameters for T2D patients: “Age”, “Sex”, “BMI”, “Duration_T2D (years)”, “HDL (mmol/L)”, “PCS”, and “WHtR”, with the “PCS” value calculable via the PCS_Calculation function in the package (details on these inputs are available in Table 1). To demonstrate, we create eight random samples based on the range of these parameters in raw data.

**Table 1:**
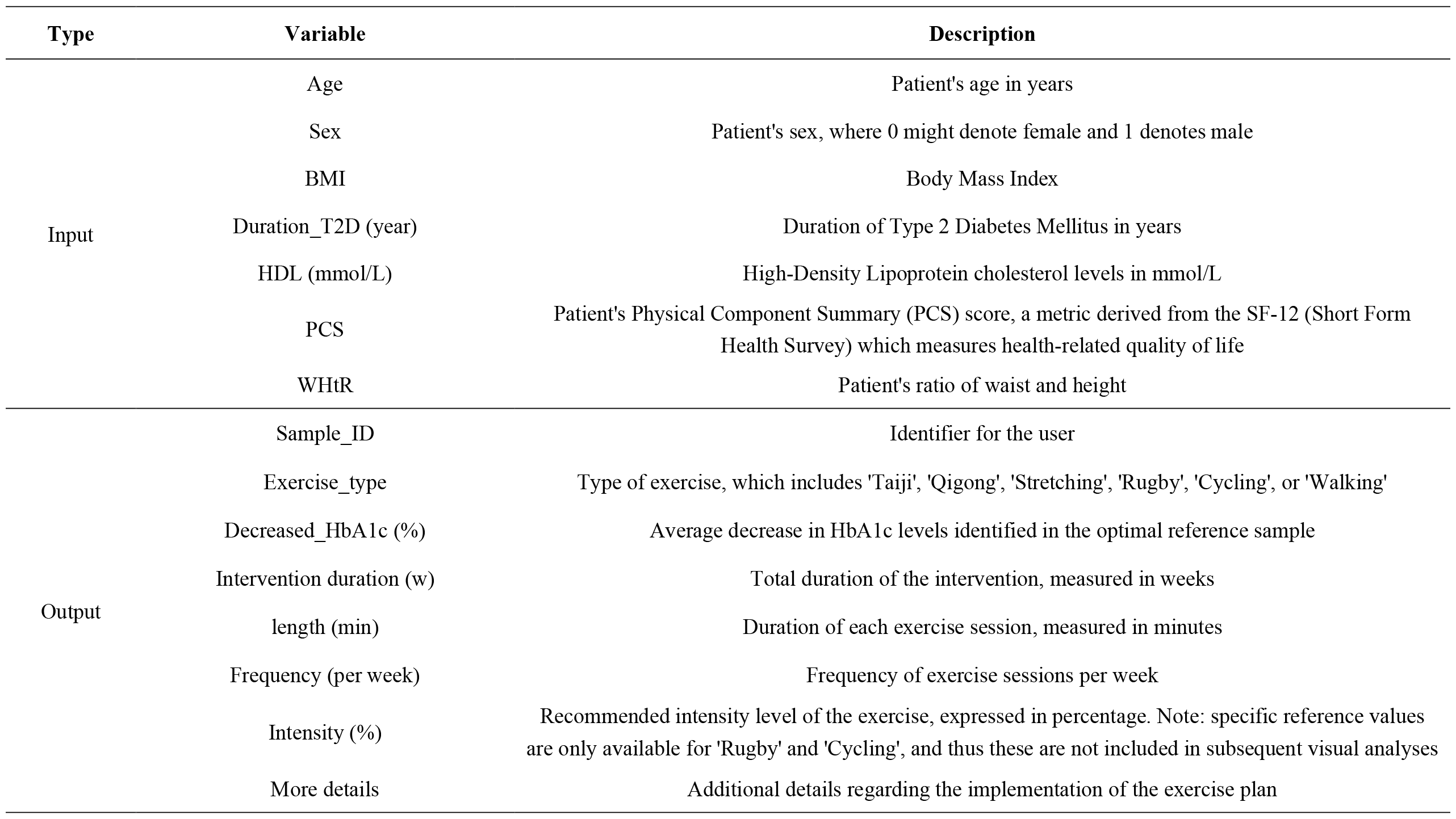
Description for input’s and output’s variables.

Upon utilizing the TailorExercisePlan function, an analysis is conducted to assess the suitability between the T2D patient’s profile and six predefined exercise types: “Taiji”, “Qigong”, “Stretching”, “Rugby”, “Cycling”, and “Walking” (See all the output parameters in Table 1). We identify and recommend exercise plans with a predicted reduction in HbA1c levels post-exercise intervention. Upon the execution of this recommendation criterion, subsequent visualization analysis is conducted with the aid of the VisualizeTailoredExercisePlan function, showcasing the outcomes for all above samples in Supplementary Figure 1.

Focusing on the sample “Xiaoli” (as depicted in Figure 1), we find that five exercise plans including Taiji, Stretching, Rugby, Cycling, and Walking, are recommended. Notably, Taiji and Stretching are forecasted to have the most substantial effect in lowering HbA1c levels, approximately by 1.2%. However, the difference in their recommendation scores, which reflect the proportion of responders, individuals who exhibit a reduction in HbA1c post-exercise intervention, in the nearest neighbor samples, is significant, being 1 and 0.6 respectively. In our analysis, the predicted fluctuation in HbA1c after each exercise plan, such as Stretching, is based on the average fluctuation observed in nearest neighbor samples. Therefore, despite having a lower recommendation score, the predicted reduction in HbA1c from Stretching exercises is similar to that from Taiji. This phenomenon might be attributable to the selection of optimal reference samples for Stretching, where responders demonstrate significantly higher positive reductions in HbA1c.

**Figure 1:**
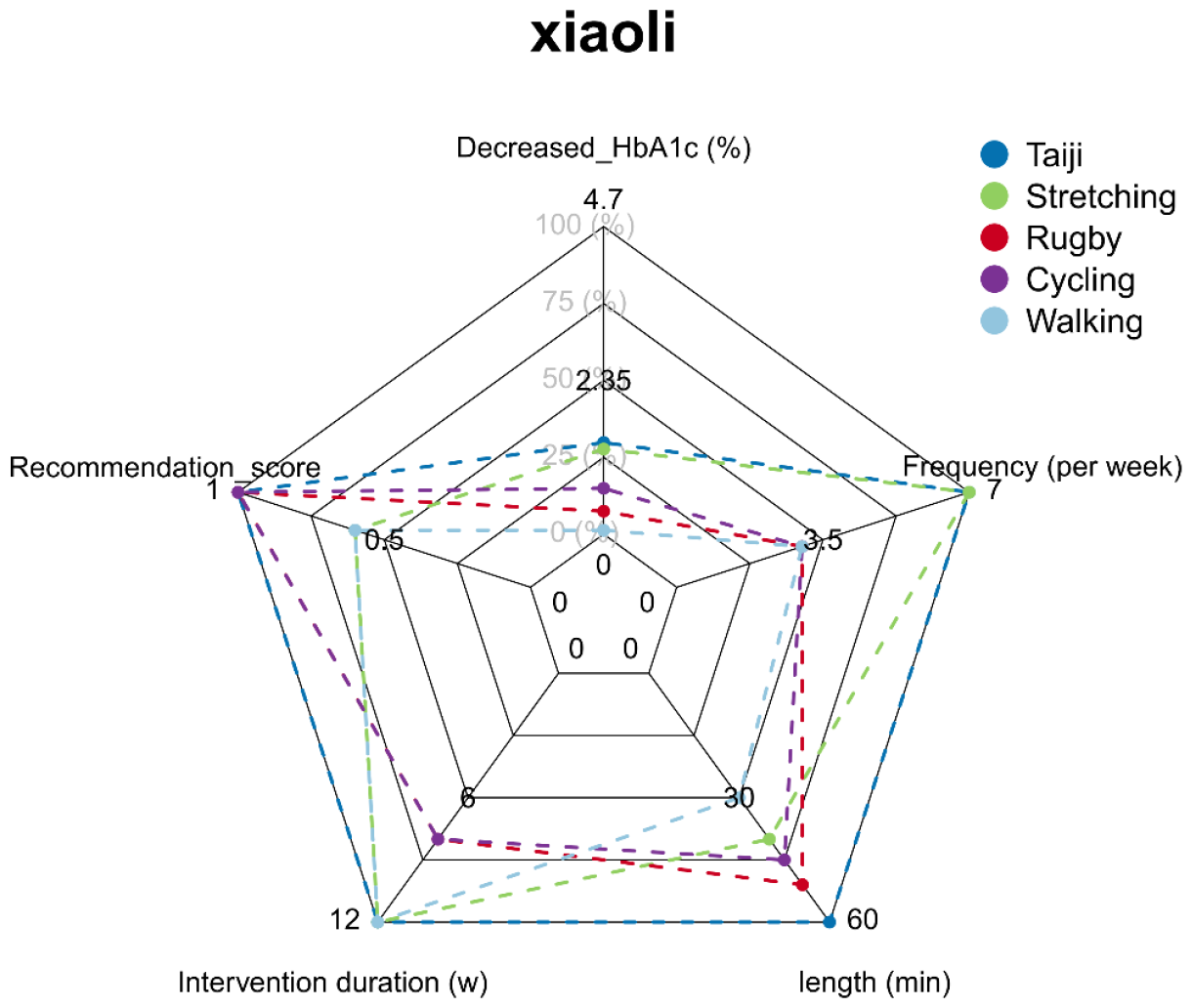
Visualization of recommended exercise plans for “Xiaoli”.

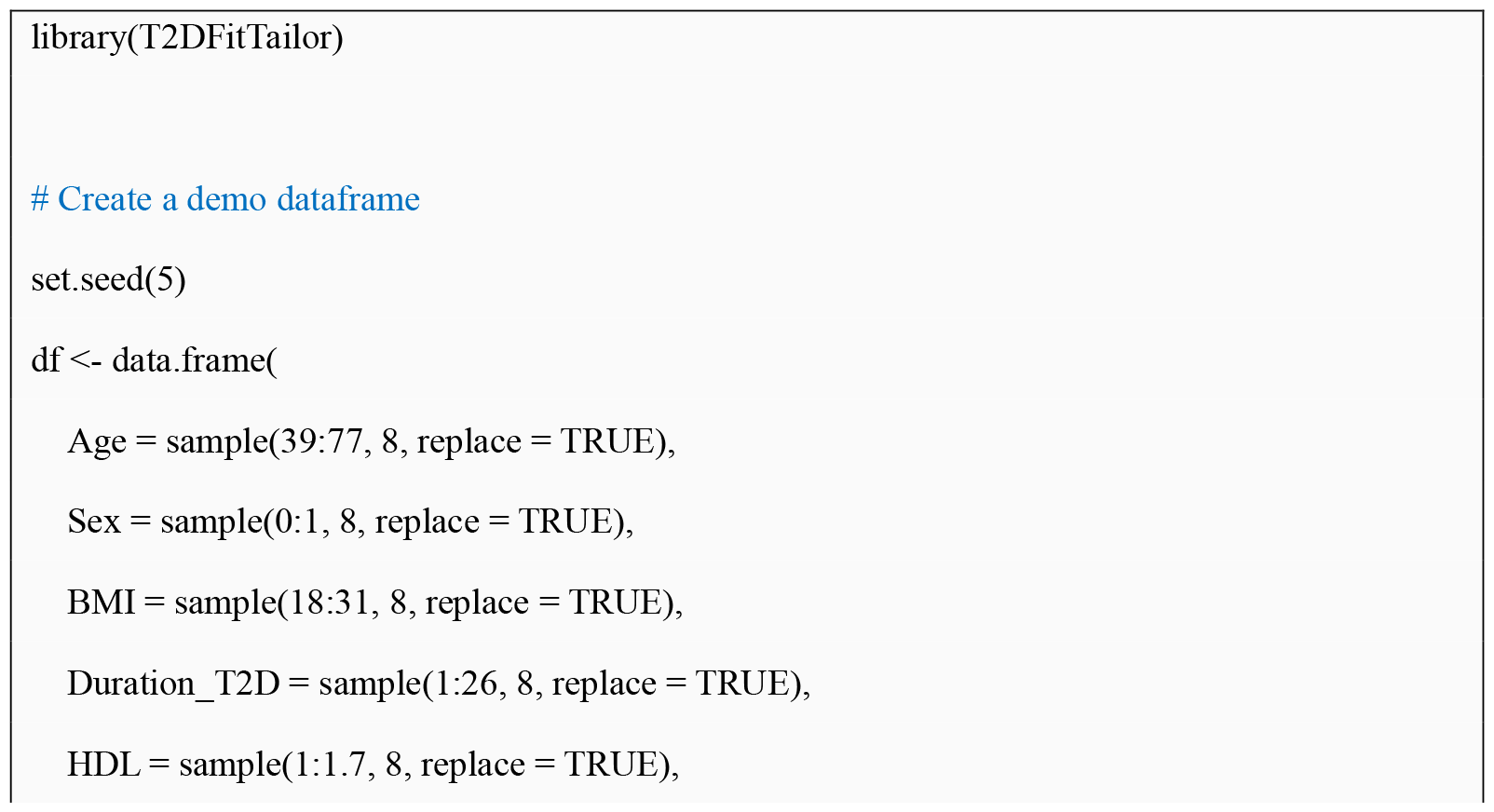
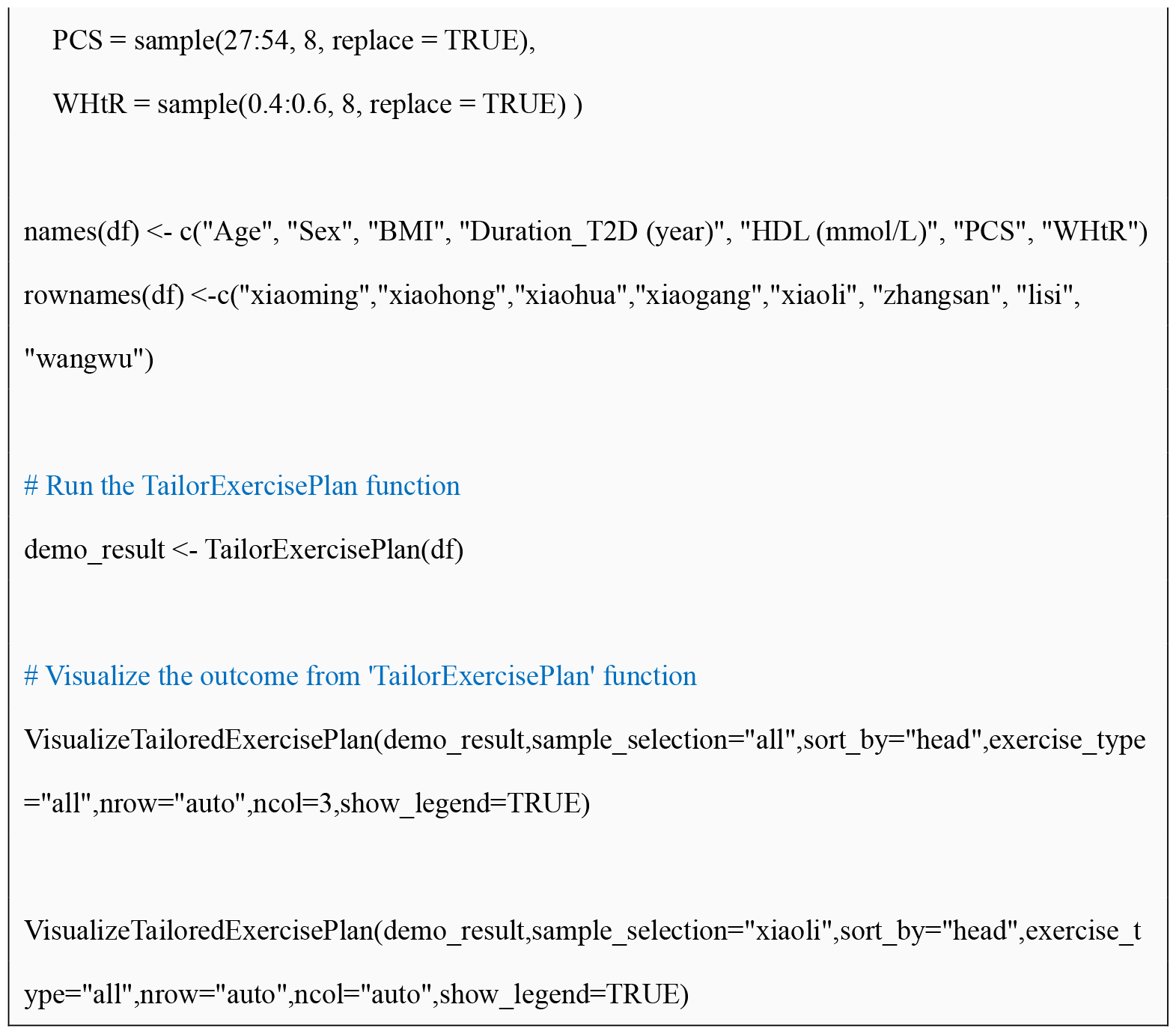

### Identification of feature panel for each exercise type

To select features meticulously enhances model generalizability and user-friendliness for T2D patients. Our initial step involved conducting an extensive literature review to identify variables associated with different forms of exercise.^(11),(12),(13)^ Subsequently, employing a 70-30 split strategy, we divided the entire dataset into training and test sets, ensuring a robust framework for model development and validation. Within this structured approach, we applied the LASSO technique to the training set to refine our variable selection, focusing on those with non-zero coefficients (Supplementary Table 1.1).

Figure 2 illustrates the finalized comprehensive feature panel, which includes seven critical variables: Age, Sex, BMI, Duration_T2D (years), HDL (mmol/L), PCS, and WHtR. Among these, Age, previously reported as a significant variable for adjusting exercise dosing, ^(14)^ emerges as the most frequently occurring variable across various exercise types, highlighting its critical role in tailoring personalized exercise plans for T2D patients.

**Figure 2:**
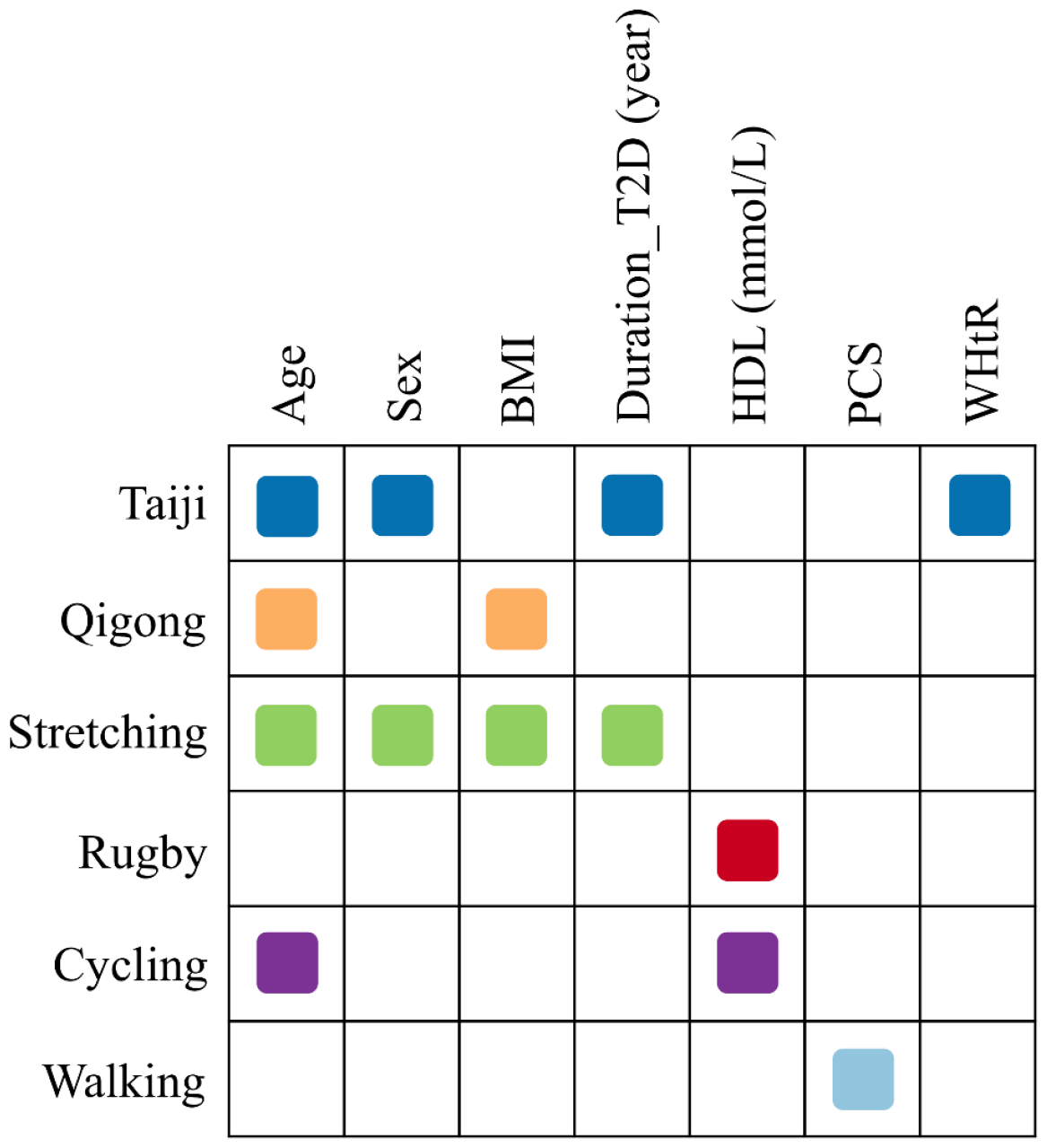
The finalized feature panel.

### Nearest neighbor-based predictions for HbA1c fluctuation post-exercise intervention

Following the construction of the feature panel, we aimed to evaluate the effects of various exercise plans on individuals with Type 2 Diabetes (T2D) using the k-Nearest Neighbors (kNN) algorithm. Initially, we determined the optimal k-value for each exercise plan based on the training set, employing a Leave-One-Out strategy. This critical step facilitates the TailorExercisePlan function to identify the k most similar reference samples to a given patient, using the average fluctuation in HbA1c values post-exercise intervention as a reference for potential outcomes. Additionally, based on exercise plans detailed in the literature from which our raw data were derived, specific exercise plans and predicted reductions in HbA1c are provided.

To confirm the effectiveness of our comprehensive exercise recommendation framework, we utilized calibration curves. Analysis depicted in Figure 3 reveals a significant linear correlation between predicted and actual HbA1c reduction values across both the training and preserved test sets (R^2^ = 0.28, MAD = 0.79, p = 5.29e-8 in the training set; R^2^ = 0.29, MAD = 0.89, p = 2.09e-04 in the test set), solidifying the framework’s predictive accuracy.

**Figure 3:**
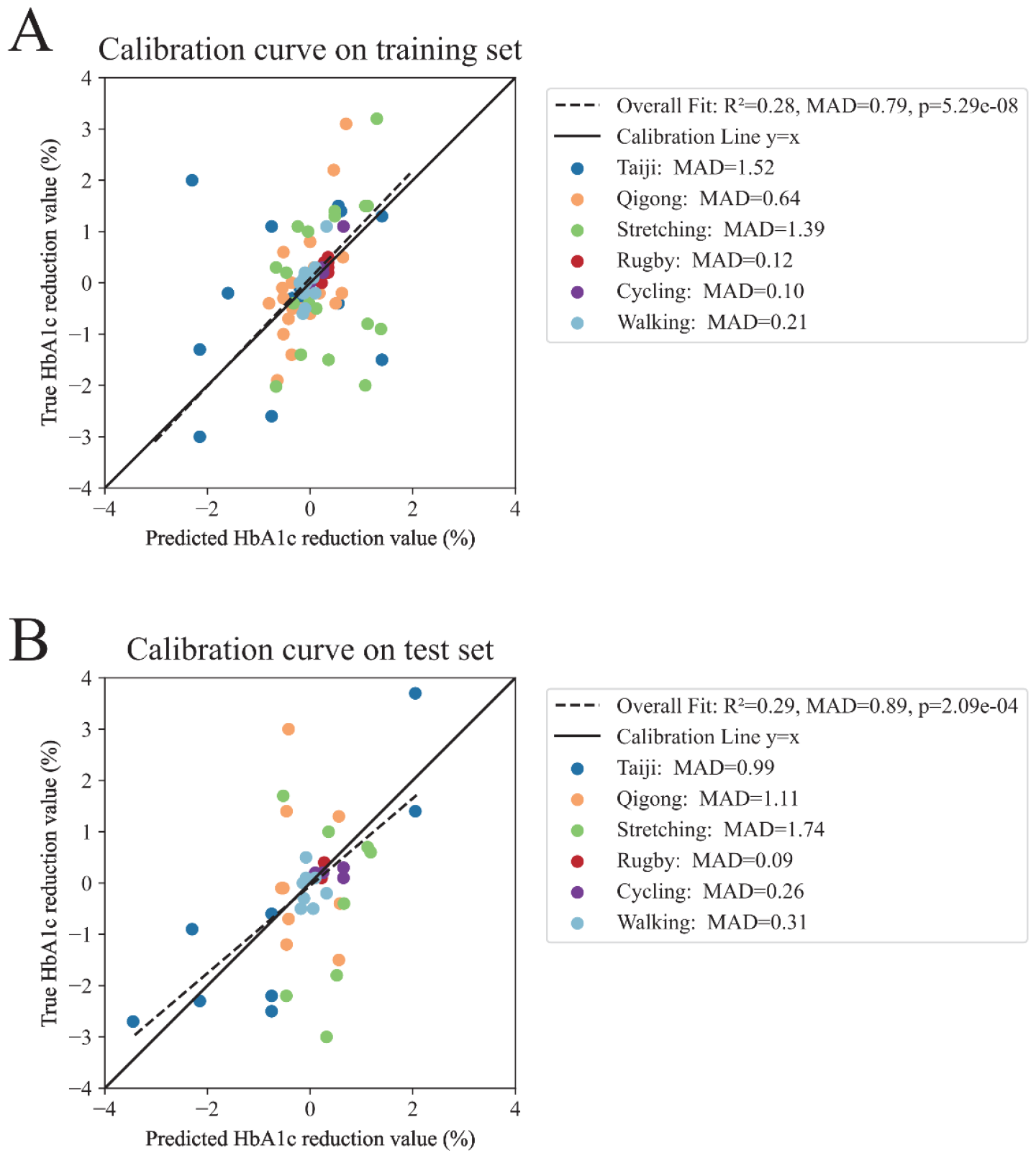
Calibration curves on (A) training set and (B) test set.

These findings collectively affirm the robustness and efficacy of the T2DFitTailor framework in tailoring exercise plans for T2D management. Through a strategic blend of feature panel selection, nearest neighbor-based predictions, and rigorous validation, we demonstrate the potential of a well-structured model to not only predict HbA1c reductions effectively but also to tailor exercise recommendations that resonate with the dynamic needs of T2D patients, thereby enhancing both the practicality and accessibility of diabetes care.

## Discussion

In this study, we advanced personalized management of Type 2 Diabetes (T2D) with the T2DFitTailor package, an innovative tool designed to tailor exercise plans to individual patient profiles, potentially addressing the challenge of varying exercise responses in T2D management. It underscores personalized healthcare’s critical role in complex diseases like T2D, with varied impacts on individuals and society. Demonstrated by cases like ‘Xiaoli’, T2DFitTailor’s potential in enhancing glycemic control through individualized exercise recommendations is evident. The integration of methods like LASSO regression for feature selection, LOO and kNN regression algorithm for predictive accuracy enhances our tool’s utility, allowing for the alignment of exercise plans with patient-specific profile and outcome prediction. T2DFitTailor sets a new precedent in diabetes care, showcasing the synergy of clinical insights and advanced data analytics to improve health outcomes.

Our study introduces the T2DFitTailor package, a advancement in tailoring exercise plans for Type 2 Diabetes (T2D) management, yet it acknowledges certain limitations. First, it employs HbA1c as the outcome metric for assessing the impact of exercise intervention. Although HbA1c has been internationally recognized as a benchmark for assessing the severity of T2D,^(15),(16),(17)^ the complexity of T2D suggests that incorporating a broader array of physiological and metabolic markers could enrich future analyses, offering a more comprehensive view of exercise benefits, such as fasting plasma glucose, C-peptide, fasting insulin levels, insulin resistance (HOMA-IR) and beta-cell function. Second, despite utilizing a train-test split strategy to validate our model and achieving a sample-to-variable ratio exceeding the minimal recommendation value of five (Supplementary Table 1.2),^(18)^ which both contribute to demonstrating the model’s strong predictive capabilities, our study faces limitations regarding independent validation of our findings. This challenge arises from the reliance on public data for studying exercise plans, making it difficult to have access to published separate papers on identical exercise plans due to lack of perceived novelty.

Moving forward, our plans for enhancing the T2DFitTailor package and its utility in Type 2 Diabetes (T2D) management include:

1. Expanding our exercise plan database through active collaborations. This will allow us to offer a more diverse range of personalized exercise plans, better meeting the varied needs of T2D patients.
2. Conducting our own T2D exercise intervention cohort study. The data collected will enrich the T2DFitTailor package with new insights and evidence, further refining its recommendations for more effective diabetes management.

## Data Availability

All data produced in the present work are contained in the manuscript

