## Supplementary figures and images for "T2DFitTailor: A tool for type 2 diabetes patients to tailor exercise plan"

### Supplementary Figure 1

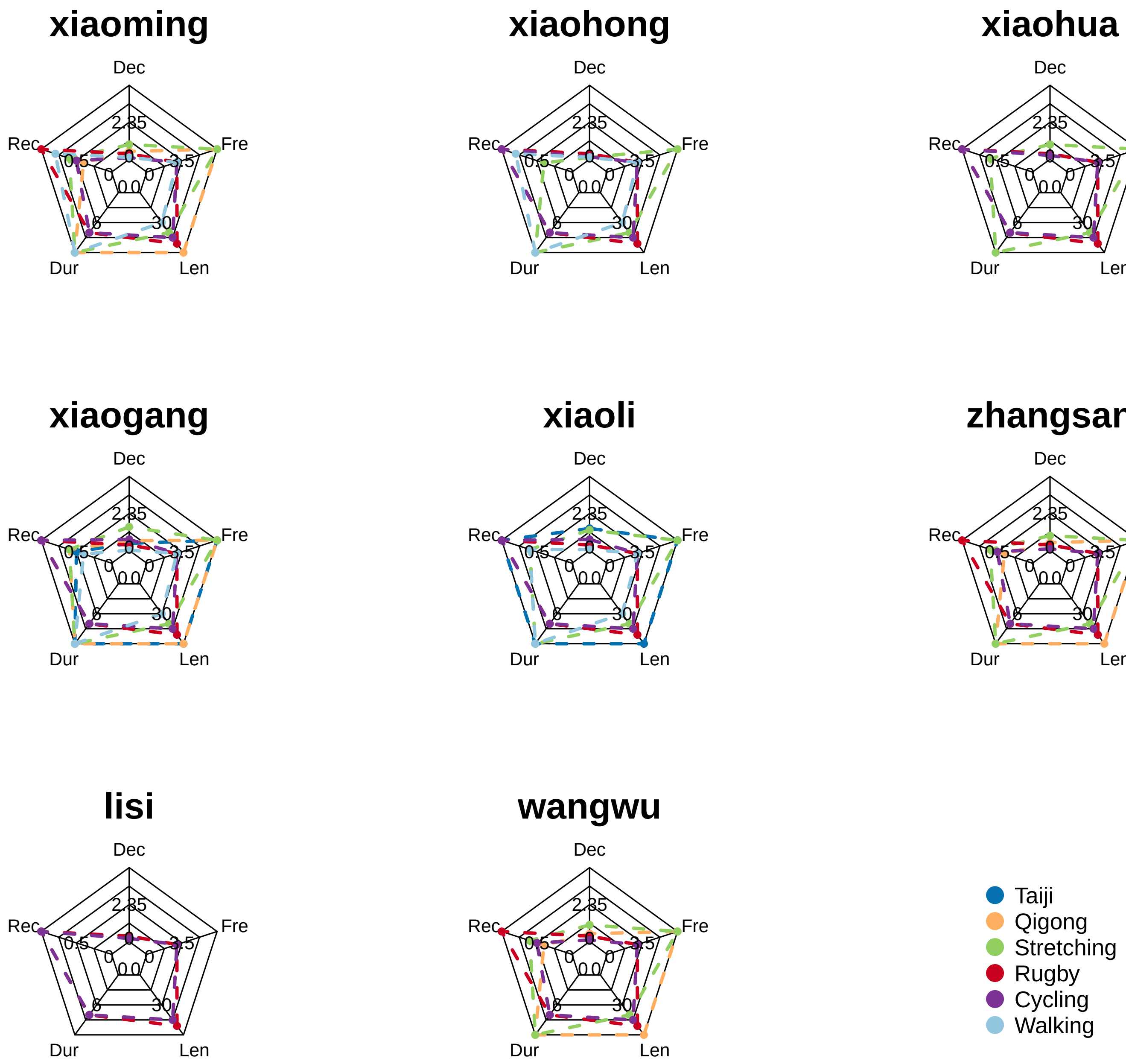
